# Comparison of the accuracy of 9 intraocular lens power calculation formulas using partial coherence interferometry

**DOI:** 10.1101/2022.04.13.22273856

**Authors:** Anthony Maroun, Mohamad El Shami, Sandra Hoyek, Joelle Antoun

## Abstract

**Purpose:** To compare the accuracy of 9 intraocular lens (IOL) power calculation formulas (SRK/T, Hoffer Q, Holladay 1, Haigis, Barrett Universal II, Kane, EVO 2.0, Ladas Super formula and Hill-RBF 3.0) using partial coherence interferometry (PCI).

**Methods:** Data from patients having uncomplicated cataract surgery with the insertion of 1 of 3 IOL types were included. All preoperative biometric measurements were performed using PCI. Prediction errors (PE) were deduced from refractive outcomes evaluated 3 months after surgery. The mean prediction error (ME), mean absolute prediction error (MAE), median absolute prediction error (MedAE), and standard deviation of prediction error (SD) were calculated, as well as the percentage of eyes with a PE within ±0.25, ±0.50, ±0.75 and ±1.00D for each formula.

**Results:** Included in the study were 126 eyes of 126 patients. Kane achieved the lowest MAE and SD across the entire sample as well as the highest percentage of PE within ±0.50D, and was proven to be more accurate than Haigis and Hoffer Q (P <.001). For an axial length of more than 26.0 mm, EVO 2.0 and Barrett obtained the lowest MAEs, with EVO 2.0 and Kane showing a higher percentage of prediction at ±0.50D compared to old generation formulas except for SRK/T (P =.04).

**Conclusion:** All investigated formulas achieved good results; there was a tendency towards better outcomes with new generation formulas, especially in atypical eyes.

## Introduction

Cataract surgery is the most frequently performed procedure in the world, with more than 30 million surgeries annually[1]. With precise biometric measures and refinement of surgical techniques, postoperative emmetropia is becoming an achievable and expected goal. Several intraocular lens power calculation formulas have been developed over the years to improve the accuracy of IOL power predictions. These formulas can be classified as either regression, vergence, artificial intelligence-based (AI) or a combination approach. Regression formulas are established retrospectively from data such as keratometry, axial length, implant power, and postoperative refraction. The calculation using regression aims to establish an equation that expresses the power of the implant as a function of these variables and, therefore, does not rely on theoretical optics. Contrarily, vergence or theoretical formulas use Gaussian optics to determine the IOL power. SRK/T, Haigis, Hoffer Q and Holladay 1 are all based on a theoretical approach and are classified as old generation formulas. On the other hand, Kane is a new formula that combines theoretical optics with both regression and AI components. It has been shown to be the most accurate formula when using optical low-coherence reflectometry (OLCR), partial coherence interferometry (PCI) or swept-source optical coherence tomography (SS-OCT)[2–7]. Another popular choice among surgeons is Barrett Universal II. Based on theoretical optics, this formula has outperformed early-generation formulas[6, 8, 9](SRK/T, Hoffer Q, Holladay 1, and Haigis), and is considered as the second-best formula[10]. Emmetropia verifying optical (EVO 2.0) is also a promising choice and is achieving good results with newer optical biometers, however, few studies have evaluated its accuracy with PCI[3, 11, 12]. Finally, Hill-RBF employs pattern recognition by artificial intelligence and was proven to be less accurate when compared to other newer generation formulas[2, 7, 10, 12]. However, because this formula is based on pattern recognition and adaptive learning, its precision is expected to increase with more data being fit into the artificial intelligence model. Very recently, a new version of this formula named Hill-RBF 3.0 was released and the database has been further increased and refined.

Our study aims to evaluate and compare the precision of traditional formulas based on vergence, with new-generation formulas, using partial coherence interferometry. In addition, we want to report the first results obtained with the new version of Hill-RBF. To the best of our knowledge, this is the first study to compare this combination of formulas using partial coherence interferometric biometry.

## Methods

The current study was conducted with the approval of the Ethics Board at Saint-Joseph University (USJ), Beirut, and adhered to the ethical principles of the Declaration of Helsinki.

A retrospective chart review was conducted on all cataract surgeries performed by one surgeon (A.J.) from January 2019 until January 2020 at Beirut Eye and Specialist Hospital (BESH), Lebanon. Patients were excluded for the following conditions: prior corneal or intraocular surgery, ocular disease, astigmatism greater than 4.00D, intra-operative or post-operative complication, postoperative corrected distance visual acuity worse than 20/40. If a patient was operated for both eyes, only the first one was included in the study to avoid bias in the correction used for the second eye. A total of 126 eyes were enrolled in this study.

All preoperative biometric measurements were made using partial coherence interferometry (IOLMaster 500, Carl Zeiss-Meditec). Phacoemulsification was performed by one surgeon through a 2.5 mm temporal incision with the insertion of one of the three following IOL models: RayOne Aspheric 600C, Superflex Aspheric 920H, C-flex Aspheric 970 C (Rayner Intraocular Lenses Ltd.). User Group for Laser Interference Biometry (ULIB) IOL constants of each model were used in the calculation. Post operative subjective spherocylindrical refractive error was assessed by optometrists 3 months after surgery and only patients with a visual acuity of 20/40 or better were enrolled.

The Hoffer Q, SRK-T, Haigis and Holladay 1 formulas were calculated using the IOLMaster 500 biometer. The remaining formulas were accessed in December 2020 from their respective web sites and ULIB SRK/T A constant for each IOL was used: Barrett Universal II (BUII)^A^, Hill-RBF recently updated to version 3.0^B^, Ladas Super Formula AI (LSF)^C^, Kane^D^ and EVO version 2.0^E^. The parameters included in the calculation were the following: axial length (AL), anterior chamber depth (ACD), keratometry (K), and patient gender. In our study, no optional variables were used (central corneal thickness, lens thickness or white-to-white). All “out-of-bounds” cases seen with Hill-RBF 3.0 were included in the study to provide a fair comparison.

## Outcome Measurements

The prediction error (PE) was calculated as the actual post-operative refraction in spherical equivalent (SE) determined 3 months post-operatively minus the predicted refraction by each formula for the power of the implanted IOL. The mean prediction error (ME), mean absolute prediction error (MAE), median absolute prediction error (MedAE), and standard deviation of prediction error (SD) were calculated, as well as the percentage of eyes with a PE ±0.25, ±0.50, ±0.75 and ±1.00D for each formula. To minimize the impact of the use of multiple IOL constants in the study, the mean error (ME) was fully “zeroed out” by adjusting the refractive prediction error for each eye by an amount equal to the overall mean prediction error for that formula as suggested by Wang et al[13].

## Statistical Analysis

Statistical analysis was performed using SPSS software (v. 26.0, IBM Corporation). Data distribution was assessed with the Kolmogorov-Smirnov test (which revealed a Gaussian distribution for the arithmetic PEs) followed by one sample t-test to evaluate whether the mean refractive error for each formula was significantly different from zero.

The comparison of absolute errors between formulas was conducted with Friedman’s test followed by post-hoc analysis using the Dunn test with Bonferroni correction for multiple comparisons, as suggested by Aristodemou et al[14]. Cochran’s Q test was used to compare the percentage of eyes within ±0.25, ±0.50, ±0.75, and ±1.00D of the predicted refraction, followed by Dunn’s post-hoc test. A *P* value of less than 0.05 was considered statistically significant.

## Results

In total, 126 eyes (69 right eyes, 54.76%; 57 left eyes, 45.24%) of 126 patients were enrolled. The demographics and biometric data of the study population are presented in Table 1. Of the 126 eyes, 101, 16 and 9 eyes were respectively implanted with the RayOne Aspheric 600C, Superflex Aspheric 920H and C-flex Aspheric 970 C IOLs. Based on AL, 12 eyes (9.5%) were classified as short (AL≤22.0 mm), 106 (84.1%) as medium (22.0<AL<26.0 mm) and 8 (6.3%) as long (AL≥26.0 mm). All formulas achieved a mean error close to zero when ULIB IOL constants were used. However, as stated before, the ME was then “zeroed out” to eliminate any systematic error. The parametric one sample t-test showed no statistically significant difference between the ME of the different formulas (*P* > .99).

**Table 1.** Demographics and biometric data

| **Parameter** | **Mean** | **SD** | **Min** | **Max** |
| --- | --- | --- | --- | --- |
| Gender | 58 | - | - | - |
| Age | 70.70 | 8.89 | 49 | 92 |
| AL (mm) | 23.63 | 1.38 | 20.73 | 30.25 |
| ACD (mm) | 3.06 | 0.45 | 2.07 | 4.25 |
| Keratometry (D) | 43.83 | 1.29 | 40.35 | 48.23 |
| IOL power (D) | 20.71 | 3.76 | 4 | 28 |

ACD = anterior chamber depth; AL = axial length; D = diopters

Table 2 shows MEs ± SD, MAEs, MedAEs, maximum absolute values, and percentages of eyes within prediction errors of ±0.25, ±0.50, ±0.75 and ±1.00D in the whole sample. Over the entire AL, Friedman’s test revealed a significant difference between formulas’ absolute prediction errors (*P* = .02). The lowest MAE value was achieved by Kane (0.299D) followed by EVO 2.0 (0.304D) and BUII (0.305D). Kane, BUII and EVO 2.0 also had the lowest SD between all formulas. On the other hand, old generation formulas gave the highest MAE and SD. For instance, the highest MAEs were achieved by Haigis (0.358D) and Hoffer Q (0.352D).

**Table 2.**
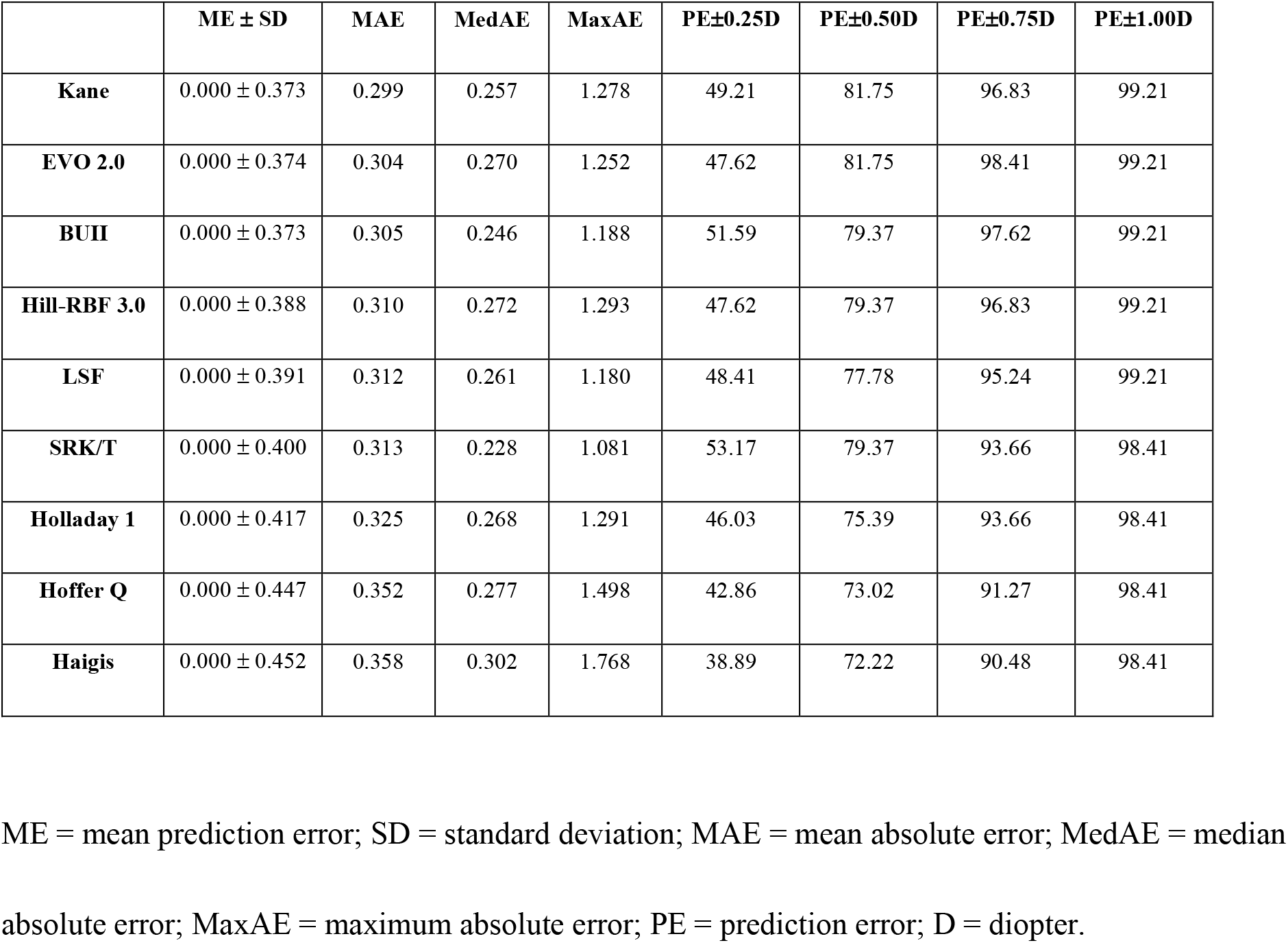
Prediction errors for each formula in the whole sample, ranked according to their respective MAE (n = 126).

| | ME ± SD | MAE | MedAE | MaxAE | PE±0.25D | PE±0.50D | PE±0.75D | PE±1.00D |
| --- | --- | --- | --- | --- | --- | --- | --- | --- |
| **Kane** | 0.000 ± 0.373 | 0.299 | 0.257 | 1.278 | 49.21 | 81.75 | 96.83 | 99.21 |
| **EVO 2.0** | 0.000 ± 0.374 | 0.304 | 0.270 | 1.252 | 47.62 | 81.75 | 98.41 | 99.21 |
| **BUII** | 0.000 ± 0.373 | 0.305 | 0.246 | 1.188 | 51.59 | 79.37 | 97.62 | 99.21 |
| **Hill-RBF 3.0** | 0.000 ± 0.388 | 0.310 | 0.272 | 1.293 | 47.62 | 79.37 | 96.83 | 99.21 |
| **LSF** | 0.000 ± 0.391 | 0.312 | 0.261 | 1.180 | 48.41 | 77.78 | 95.24 | 99.21 |
| **SRK/T** | 0.000 ± 0.400 | 0.313 | 0.228 | 1.081 | 53.17 | 79.37 | 93.66 | 98.41 |
| **Holladay 1** | 0.000 ± 0.417 | 0.325 | 0.268 | 1.291 | 46.03 | 75.39 | 93.66 | 98.41 |
| **Hoffer Q** | 0.000 ± 0.447 | 0.352 | 0.277 | 1.498 | 42.86 | 73.02 | 91.27 | 98.41 |
| **Haigis** | 0.000 ± 0.452 | 0.358 | 0.302 | 1.768 | 38.89 | 72.22 | 90.48 | 98.41 |

ME = mean prediction error; SD = standard deviation; MAE = mean absolute error; MedAE = median absolute error; MaxAE = maximum absolute error; PE = prediction error; D = diopter.

Interestingly, the lowest MedAEs were obtained with SRK/T (0.228D), BUII (0.246D) and Kane (0.257D) while Haigis and Hoffer Q had the highest MedAEs. Dunn post-test analysis showed that only the following paired comparison had statistically significant differences (*P* < .001): Kane vs Haigis and Kane vs Hoffer Q.

According to Cochran’s Q test, the proportion of eyes with a PE within ±0.25, ±0.50, and ±0.75D, displayed in figure 1, were statistically different among the investigated formulas (*P* = .03 for ±0.25D, *P* = .01 for ±0.50D and *P* < .001 for ±0.75D). SRK/T achieved the highest percentage of PE within ±0.25D (53.17%) followed by BUII (51.59%) and Kane (49.21%). Dunn’s post-test analysis with Bonferroni correction showed significant difference only in SRK/T vs Haigis (*P* = .03) in the ±0.25D category. Every formula, except for Haigis and Hoffer Q, yielded a percentage of eyes within ±0.50D higher than 75%. No significant difference was found in pairwise comparisons. Moreover, all formulas had a percentage of 90% or higher within the ±0.75D interval. Within this range, old generation formulas ranked last, having the lowest percentage. Whereas EVO 2.0 achieved the highest percentage (98.41%), showing significant difference both with Haigis (*P* = .01) and Hoffer Q (*P* = .04). In addition, BUII had the second highest percentage at 97.62% and was significantly superior to Haigis (*P* = .04).

**Figure 1.**
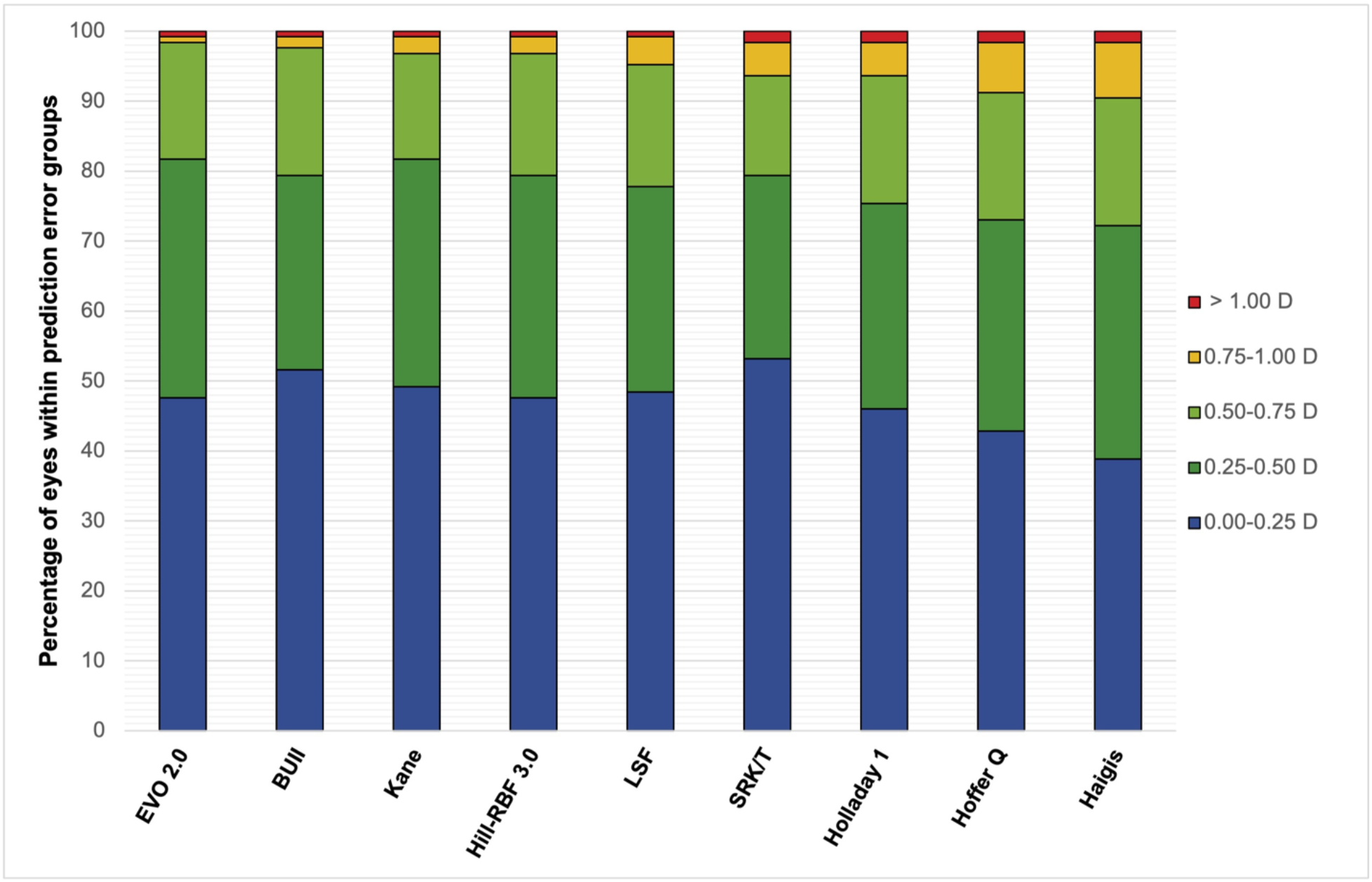
Stacked histogram, showing the percentage of eyes within ±0.25, ±0.50, ±0.75, ±1.00 and >1.00D of the predicted refraction, in the whole sample.

## Results by Axial Length

The MEs ± SD, MAEs, MedAEs, maximum values, and percentages of eyes within prediction errors of ±0.25, ±0.50, ±0.75 and ±1.00D for the subgroup of short eyes (AL≤22.0 mm; n=12) and long eyes (AL≥26.0 mm; n=8) are shown respectively in table 3 and table 4. For short eyes, Friedman’s test showed no significant differences between all formulas. However, it is worth to point out that Kane had the lowest MAE (0.249D), lowest SD (0.278), and second lowest MedAE (0.216D). No significant difference was seen between all formulas when comparing the proportion of eyes with a PE within ±0.25D, ±0.50D, ±0.75D and ±1.00D.

**Table 3.** Prediction errors for each formula in short eyes, ranked according to their respective MAE (n = 12).

| | ME ± SD | MAE | MedAE | MaxAE | PE±0.25D | PE±0.50D | PE±0.75D | PE±1.00D |
| --- | --- | --- | --- | --- | --- | --- | --- | --- |
| **Kane** | 0.028 ± 0.278 | 0.249 | 0.216 | 0.435 | 66.67 | 91.67 | 91.67 | 91.67 |
| **EVO 2.0** | -0.003 ± 0.281 | 0.249 | 0.241 | 0.419 | 50 | 91.67 | 91.67 | 91.67 |
| **Holladay 1** | 0.022 ± 0.332 | 0.270 | 0.227 | 0.589 | 58.33 | 83.33 | 91.67 | 91.67 |
| **Hoffer Q** | -0.144 ± 0.350 | 0.291 | 0.322 | 0.710 | 41.67 | 75 | 91.67 | 91.67 |
| **Haigis** | -0.130 ± 0.374 | 0.293 | 0.250 | 0.814 | 50.0 | 75 | 83.33 | 91.67 |
| **SRK/T** | 0.088 ± 0.356 | 0.298 | 0.225 | 0.713 | 58.33 | 75.0 | 91.67 | 91.67 |
| **BUII** | -0.001 ± 0.339 | 0.300 | 0.249 | 0.591 | 50.0 | 75 | 91.67 | 91.67 |
| **Hill-RBF 3.0** | -0.059 ± 0.394 | 0.303 | 0.252 | 0.990 | 50.0 | 83.33 | 83.33 | 91.67 |
| **LSF** | -0.086 ± 0.409 | 0.316 | 0.193 | 0.901 | 58.33 | 75 | 83.33 | 91.67 |

ME = mean prediction error; SD = standard deviation; MAE = mean absolute error; MedAE = median absolute error; MaxAE = maximum absolute error; PE = prediction error; D = diopter.

**Table 4.** Prediction errors for each formula in long eyes, ranked according to their respective MAE (n = 8).

|  | **ME ± SD** | **MAE** | **MedAE** | **MaxAE** | **PE±0.25D** | **PE±0.50D** | **PE±0.75D** | **PE±1.00D** |
| --- | --- | --- | --- | --- | --- | --- | --- | --- |
| **EVO 2.0** | 0.087 ± 0.278 | 0.240 | 0.190 | 0.431 | 62.5 | 100.0 | 100.0 | 100.0 |
| **BUII** | 0.123 ± 0.298 | 0.244 | 0.227 | 0.526 | 50.0 | 87.50 | 100.0 | 100.0 |
| **Kane** | 0.097 ± 0.326 | 0.266 | 0.256 | 0.475 | 50.0 | 100.0 | 100.0 | 100.0 |
| **LSF** | 0.199 ± 0.280 | 0.267 | 0.198 | 0.539 | 62.5 | 87.5 | 100.0 | 100.0 |
| **Hill-RBF 3.0** | 0.318 ± 0.263 | 0.318 | 0.282 | 0.632 | 50.0 | 62.50 | 100.0 | 100.0 |
| **SRK/T** | 0.165 ± 0.412 | 0.340 | 0.265 | 0.904 | 50.0 | 87.50 | 87.50 | 100.0 |
| **Haigis** | 0.497 ± 0.191 | 0.497 | 0.469 | 0.786 | 50.0 | 50.0 | 75.0 | 100.0 |
| **Holladay 1** | 0.513 ± 0.385 | 0.523 | 0.606 | 1.099 | 25.0 | 37.50 | 87.50 | 87.50 |
| **Hoffer Q** | 0.587 ± 0.290 | 0.587 | 0.587 | 0.895 | 12.50 | 37.50 | 62.50 | 100.0 |

ME = mean prediction error; SD = standard deviation; MAE = mean absolute error; MedAE = median absolute error; MaxAE = maximum absolute error; PE = prediction error; D = diopter.

In long eyes, significant difference between the formulas’ absolute prediction errors was found (*P* < .001). New generation formulas ranked first, having the lowest MAE, whereas old generation formulas including SRK/T ranked last. In fact, EVO 2.0, BUII and Kane yielded the lowest MAE and MedAE. In post-hoc analysis, Hoffer Q produced a higher absolute error when compared to EVO 2.0, BUII and Kane (*P* < .001).

According to the Cochran Q test, the proportion of eyes with a PE within ±0.25D and ±0.50D were significantly different (*P* = .01 for ±0.25D and *P* < 0.001 for ±0.5D). EVO 2.0 and LSF provided the highest prediction within ±0.25D, however no significant difference in pairwise comparison was seen. Regarding the proportion of eyes within ±0.50D, EVO 2.0 and Kane gave rise to the highest percentage followed by BUII, LSF and SRK/T. In addition, Hoffer Q and Holladay 1 had a lower percentage than Kane and EVO 2.0 (*P* = .04). Finally, all newer formulas had no absolute prediction error higher than 0.75D in this subgroup. On the other hand, all old generation formulas including SRK/T had PEs higher than 0.75D.

Regarding the new version of Hill-RBF, 4 (3.2%) “out of bounds” values appeared in the whole sample. No statistical significance was seen when comparing Hill-RBF 3.0 to old generation or other new generation formulas. In the whole sample, this formula ranked fourth for the lowest MAE, SD and percentage of eyes with a PE ±0.75D. In short eyes, the accuracy of Hill-RBF decreased, achieving the second highest MAE (0.303D) and MedAE (0.252D). On the other hand, in long eyes, Hill-RBF improved its performance and yielded a lower MAE (0.318D) and MedAE (0.282D) compared to old generation formulas.

## Discussion

Our data shows that good refractive outcomes can be achieved with all formulas, however, there was a tendency towards better outcomes with newer formulas. In fact, all new generation formulas had a prediction error of ±0.50D or less in at least 75% of eyes, and ranked first for the lowest MAE and SD.

Overall, EVO 2.0, Kane and BUII yielded the most accurate outcomes and we found only modest differences between these formulas. Kane had the lowest MAE and SD in the whole sample as well as the highest percentage of PE within ±0.50D and was proven to be more effective than old generation formulas like Hoffer Q and Haigis (*P* < .001). Several studies have previously reported the same finding using PCI biometers[4, 12] or newer technologies[3, 5, 11, 15]. In fact, multiple articles have recently confirmed that Kane is the most accurate formula overall and in each axial length subgroup[4, 7, 10]. In our study, Kane had also the lowest MAE, SD and highest percentage of PE within ±0.50D in the short AL subgroup. However, in the long AL subgroup it was outperformed by EVO 2.0 and BUII. Similar results were also found in previous literature[15].

The EVO 2.0 formula provided accurate outcomes across the entire sample and in all AL subgroups making it one of the most accurate formulas. In short eyes, although no statistical significance was seen, EVO 2.0 had the lowest MAE and the second lowest SD. This formula had also the highest proportion of eyes with a PE within ±0.50D and ±0.75D, in agreement with previous literature^2^. In fact, the high accuracy of EVO 2.0 in short eyes has been reported in previous larger studies with different biometers [15, 16], confirming that EVO 2.0 is more accurate than old generation formulas like Holladay 1 and Hoffer Q. Interestingly, this formula also proved to be the most accurate one in the long eyes subgroup, having the lowest MAE, MedAE and highest percentage of eyes within a PE ±0.50D. This regularity across the entire AL range, also reported by Hipólito-Fernandes et al[3], makes EVO 2.0 an adequate choice.

The Barrett Universal II was previously considered as the most accurate IOL formula[6, 8, 9]. In our study, this formula confirmed its reputation remaining one of the most accurate formulas across the whole sample and in the long eyes subgroup. However, in short eyes, Barrett showed a lower accuracy achieving one of the highest MAE, MedAE (SD) at 0.300D, 0.249D (0.339), in agreement with previous studies[3–5, 15].

Hill RBF 3.0 was released in December of 2020, after refining and increasing the size of the database. 4 (3.2%) out of bounds values were seen in the sample and were included in the study. In comparison, the Hill RBF 2.0 was found to have 11.5% out of bound values in a study by Connell et al[5].Therefore, as expected, with further increase of the database the formula is showing less out-of-bound measurements. In our study, Hill-RBF 3.0 showed a higher accuracy in comparison with older generation formulas except in short eyes, but did not outperform other new generation formulas. No published study on Hill RBF 3.0 exists and therefore we were not able to compare our results. When comparing the results seen in the current version with the results seen in the previous version in several other studies[15, 17], we found that the third version has improved its accuracy slightly when considering the whole sample. However, Kane, EVO 2.0 and BUII still yielded slightly lower MAE, SD and MedAE.

Between older formulas, SRK/T ranked first, achieving the lowest MedAE, MAE and the highest percentage (79.37%) of eyes with a PE within ±0.50D when compared to other old generations formulas. More importantly, SRK/T showed only little differences with newer formulas like LSF and Hill-RBF. In contrast, we noticed a tendency towards better results with Kane, EVO 2.0 and BUII in the whole sample and in each AL subgroup. The accuracy of the SRK/T was similar in a recent study by Savini et al[12] but better than in previous larger studies[4, 6].

Finally, Hoffer Q, Haigis and Holladay 1 formulas presented the lowest accuracy in the whole sample. Hoffer Q resulted in a MAE (MedAE) of 0.352D (0.277D) whereas Haigis achieved 0.358D (0.302D). In fact, Kane proved to be superior to both formulas (*P* < .001 for Haigis and *P* < .001 for Hoffer Q). EVO 2.0 yielded also a significantly higher percentage of PE within ±0.50D (*P* = .02 for Hoffer Q, *P* = .04 for Haigis).

On the other hand, as expected, Hoffer Q and Holladay 1 improved their accuracy in short eyes, as already proven in the literature[18, 19]. No statistical significance was seen in short eyes when comparing Hoffer Q and Holladay 1 with newer formulas. Nevertheless, we observed a lower MAE and SD with Kane and EVO 2.0. In a larger study by Darcy et al.[4], Kane was significantly more accurate in short eyes than Hoffer Q, Holladay 1 and all other old generation formulas (*P* < 0.001), proving that with bigger samples new generation formulas outperform older generation in all AL subgroups.

The limitations of our study include the small number of eyes that were enrolled (n=126), and especially in the subgroups of AL≤22.0 mm (n=12) and AL≥26.0 mm (n=8). The use of 3 different IOL models can be considered as a limitation in this study, however as stated before the ME was zeroed out to avoid any systemic error. The absence of optional variables (lens thickness, central corneal thickness and white-to-white measurements) is a third limitation. Including the optional variables in the calculation can potentially produce better outcomes. However, given the widespread use of partial coherence interferometry and older biometers, our study can help surgeon refine their choice about IOL formulas.

## Conclusion

This study shows that all formulas can be successfully used to calculate the IOL power with PCI, however, new generation formulas like Kane, EVO 2.0 and BUII can help us achieve better results. In fact, EVO 2.0 and Kane were the most accurate formulas across the entire sample and in all AL subgroups. On the other hand, BUII decreased its precision in short eyes. Finally, regarding Hill-RBF 3.0, our data suggest that this new version gives less out of bound values and is slightly more precise compared to the previous version, but Kane, Barrett and EVO 2.0 remain superior.

## Data Availability

All data produced in the present study are available upon reasonable request to the authors

## References

[1] Liu Y-C, Wilkins M, Kim T, Malyugin B, Mehta JS. Cataracts. The Lancet 2017;390(10094):600–12.

[2] Savini G, Hoffer KJ, Balducci N, Barboni P, Schiano-Lomoriello D. Comparison of formula accuracy for intraocular lens power calculation based on measurements by a swept-source optical coherence tomography optical biometer. J Cataract Refract Surg 2020;46(1):27–33.

[3] Hipolito-Fernandes D, Elisa Luis M, Gil P, Maduro V, Feijao J, Yeo TK, et al. VRF-G, a New Intraocular Lens Power Calculation Formula: A 13-Formulas Comparison Study. Clin Ophthalmol 2020;14:4395–402.

[4] Darcy K, Gunn D, Tavassoli S, Sparrow J, Kane JX. Assessment of the accuracy of new and updated intraocular lens power calculation formulas in 10 930 eyes from the UK National Health Service. J Cataract Refract Surg 2020;46(1):2–7.

[5] Connell BJ, Kane JX. Comparison of the Kane formula with existing formulas for intraocular lens power selection. BMJ Open Ophthalmol 2019;4(1):e000251.

[6] Melles RB, Holladay JT, Chang WJ. Accuracy of Intraocular Lens Calculation Formulas. Ophthalmology 2018;125(2):169–78.

[7] Melles RB, Kane JX, Olsen T, Chang WJ. Update on Intraocular Lens Calculation Formulas. Ophthalmology 2019;126(9):1334–5.

[8] Cooke DL, Cooke TL. Comparison of 9 intraocular lens power calculation formulas. J Cataract Refract Surg 2016;42(8):1157–64.

[9] Kane JX, Van Heerden A, Atik A, Petsoglou C. Intraocular lens power formula accuracy: Comparison of 7 formulas. J Cataract Refract Surg 2016;42(10):1490–500.

[10] Kane JX, Chang DF. Intraocular Lens Power Formulas, Biometry, and Intraoperative Aberrometry: A Review. Ophthalmology 2021;128(11):e94–e114.

[11] Cheng H, Kane JX, Liu L, Li J, Cheng B, Wu M. Refractive Predictability Using the IOLMaster 700 and Artificial Intelligence-Based IOL Power Formulas Compared to Standard Formulas. J Refract Surg 2020;36(7):466–72.

[12] Savini G, Di Maita M, Hoffer KJ, Naeser K, Schiano-Lomoriello D, Vagge A, et al. Comparison of 13 formulas for IOL power calculation with measurements from partial coherence interferometry. Br J Ophthalmol 2021;105(4):484–9.

[13] Wang L, Koch DD, Hill W, Abulafia A. Pursuing perfection in intraocular lens calculations: III. Criteria for analyzing outcomes. J Cataract Refract Surg 2017;43(8):999–1002.

[14] Aristodemou P, Knox Cartwright NE, Sparrow JM, Johnston RL. Statistical Analysis for Studies of Intraocular Lens Formula Accuracy. Am J Ophthalmol 2015;160(5):1085–6.

[15] Carmona-Gonzalez D, Castillo-Gomez A, Palomino-Bautista C, Romero-Dominguez M, Gutierrez-Moreno MA. Comparison of the accuracy of 11 intraocular lens power calculation formulas. Eur J Ophthalmol 2021;31(5):2370–6.

[16] Kane JX, Melles RB. Intraocular lens formula comparison in axial hyperopia with a high-power intraocular lens of 30 or more diopters. J Cataract Refract Surg 2020;46(9):1236–9.

[17] Nemeth G, Modis L, Jr. Accuracy of the Hill-radial basis function method and the Barrett Universal II formula. Eur J Ophthalmol 2021;31(2):566–71.

[18] Day AC, Foster PJ, Stevens JD. Accuracy of intraocular lens power calculations in eyes with axial length <22.00 mm. Clin Exp Ophthalmol 2012;40(9):855–62.

[19] Srivannaboon S, Chirapapaisan C, Chirapapaisan N, Lertsuwanroj B, Chongchareon M. Accuracy of Holladay 2 formula using IOLMaster parameters in the absence of lens thickness value. Graefes Arch Clin Exp Ophthalmol 2013;251(11):2563–7.

## Other cited materials

A. Barrett GD. Barrett Universal II Formula. Singapore, Asia-Pacific Association of Cataract and Refractive Surgeons. Available at: https://calc.apacrs.org/barrett_universal2105. Accessed December 2020.

B. Hill W. Hill-RBF calculator version 3.0. Available at: https://rbfcalculator.com/ online/index.html https://rbfcalculator.com/online/index.html. Accessed December 2020.

C. Ladas JG, Siddiqui AA, Devgan U, Jun AS. Ladas Super Formula AI. Available at: http://www.iolcalc.com/. Accessed December 2020.

D. Kane JX. Kane Formula. Available at: https://www.iolformula.com/. Accessed December 2020.

E. Yeo TK. EVO Formula 2.0 Calculator. Available at: https://www.evoiolcalculator.com/calculator.aspx. Accessed December 2020.

